# SARS-CoV-2 antibody magnitude and detectability are driven by disease severity, timing, and assay

**DOI:** 10.1101/2021.03.03.21251639

**Authors:** Michael J. Peluso, Saki Takahashi, Jill Hakim, J. Daniel Kelly, Leonel Torres, Nikita S. Iyer, Keirstinne Turcios, Owen Janson, Sadie E. Munter, Cassandra Thanh, Christopher C. Nixon, Rebecca Hoh, Viva Tai, Emily A. Fehrman, Yanel Hernandez, Matthew A. Spinelli, Monica Gandhi, Mary-Ann Palafox, Ana Vallari, Mary A. Rodgers, John Prostko, John Hackett, Lan Trinh, Terri Wrin, Christos J. Petroplolous, Charles Y. Chiu, Philip J. Norris, Clara DiGermanio, Mars Stone, Michael P. Busch, Susanna K. Elledge, Xin X. Zhou, James A. Wells, Albert Shu, Theodore W. Kurtz, John E. Pak, Wesley Wu, Peter D. Burbelo, Jeffrey I. Cohen, Rachel L. Rutishauser, Jeffrey N. Martin, Steven G. Deeks, Timothy J. Henrich, Isabel Rodriguez-Barraquer, Bryan Greenhouse

**Author notes:** These authors contributed equally (Michael J. Peluso, Saki Takahashi, Jill Hakim). These authors contributed equally (Timothy J. Henrich, Isabel Rodriguez-Barraquer, Bryan Greenhouse).

## Abstract

Serosurveillance studies are critical for estimating SARS-CoV-2 transmission and immunity, but interpretation of results is currently limited by poorly defined variability in the performance of antibody assays to detect seroreactivity over time in individuals with different clinical presentations. We measured longitudinal antibody responses to SARS-CoV-2 in plasma samples from a diverse cohort of 128 individuals over 160 days using 14 binding and neutralization assays. For all assays, we found a consistent and strong effect of disease severity on antibody magnitude, with fever, cough, hospitalization, and oxygen requirement explaining much of this variation. We found that binding assays measuring responses to spike protein had consistently higher correlation with neutralization than those measuring responses to nucleocapsid, regardless of assay format and sample timing. However, assays varied substantially with respect to sensitivity during early convalescence and in time to seroreversion. Variations in sensitivity and durability were particularly dramatic for individuals with mild infection, who had consistently lower antibody titers and represent the majority of the infected population, with sensitivities often differing substantially from reported test characteristics (e.g., amongst commercial assays, sensitivity at 6 months ranged from 33% for ARCHITECT IgG to 98% for VITROS Total Ig). Thus, the ability to detect previous infection by SARS-CoV-2 is highly dependent on the severity of the initial infection, timing relative to infection, and the assay used. These findings have important implications for the design and interpretation of SARS-CoV-2 serosurveillance studies.

## INTRODUCTION

Despite advances in severe acute respiratory syndrome-coronavirus-2 (SARS-CoV-2) prevention and treatment, the novel coronavirus continues to infect individuals at an unprecedented rate. Because vaccination programs remain limited in scope, millions of individuals worldwide continue to rely upon natural post-infection immunity for protection from reinfection. Serosurveillance studies measuring the prevalence of antibodies to SARS-CoV-2 have been and will continue to be a key means for estimating transmission over time and extrapolating potential levels of immunity in populations, though precise correlates of protection have yet to be established. However, limited available data on the sensitivity of antibody assays to detect prior infection - particularly in appropriately representative populations and over time - make it difficult to accurately interpret results from these studies.^1^ For these reasons, longitudinal characterization of antibody responses following SARS-CoV-2 infection with a range of clinical presentations is an important research gap and will be critical to interpreting seroepidemiological data and informing public health responses to the pandemic.

Infection with SARS-CoV-2 is associated with substantial variability in disease presentation, with severity ranging from asymptomatic infection to the need for high-level oxygen support and mechanical ventilation.^2,3^ There appear to be important relationships between the severity of illness and the magnitude and durability of the antibody response,^4–12^ but limited data are available evaluating the contributions of demographic factors and clinical features. Numerous platforms are available for the detection of antibody responses to SARS-CoV-2, which rely on different viral antigens and utilize different assay methods, and there is no guarantee that they will provide comparable data. With a few notable exceptions,^6,7^ most studies to date have produced antibody data from a single or limited number of platforms to evaluate antibody responses following infection.^8–10,13–15^ Comparisons across platforms and assay format differences (e.g., direct vs. indirect detection), including the correlation between binding assays and neutralization capacity, have thus far have been limited.^4,7,16^

Here, we characterize the antibody responses to SARS-CoV-2 among a diverse cohort of individuals with documented infection, with a focus on investigating (1) the determinants of the magnitude and durability of humoral immune responses across a spectrum of disease severity, (2) the relationship between antibody responses across a wide variety of binding assay platforms (13 total) and their correlation with neutralization capacity, and (3) the implications of individual, temporal, and assay variability for serosurveillance. Our findings provide insight into the interpretation of antibody test results, and have important implications for our understanding of humoral immunity to natural infection as well as for serosurveillance.

## METHODS

### Study cohort

Participants were volunteers in the University of California, San Francisco-based Long-term Impact of Infection with Novel Coronavirus (LIINC) natural history study (NCT04362150). All volunteers signed informed consents for the study. LIINC is an observational cohort that enrolls individuals with SARS-CoV-2 infection documented by clinical nucleic acid amplification testing who have recovered from the acute phase of infection. Volunteers are recruited by clinician- or self-referral. They are eligible to enroll between 14 and 90 days after onset of COVID-19 symptoms and are offered monthly visits until 4 months after illness onset; they are then seen every 4 months thereafter.

Clinical data from the initial LIINC study visit was used for this analysis. At this visit, LIINC participants underwent a detailed clinical interview conducted by a study physician or research coordinator using a standardized data collection instrument. Demographic data collected included age, sex, gender, race, ethnicity, education level, income level, and housing status. Data related to SARS-CoV-2 infection included the date and circumstances of diagnosis, illness, and treatment history. Each participant was asked to estimate the date of symptom onset in relation to the timing of their first SARS-CoV-2 nucleic acid amplification test result. Participants were questioned regarding the presence, duration (in days), and current status of a list of COVID-19 symptoms and additional somatic symptoms derived from the Patient Health Questionnaire ^17^, as well as measures of quality of life derived from the EQ-5D-5L Instrument ^18^. We determined from medical records whether each individual was hospitalized (defined as spending >24 hours in the emergency department or hospital) and whether they required supplemental oxygen, admission to an intensive care unit (ICU), or mechanical ventilation. Past medical history was ascertained and concomitant medications recorded.

At each visit, blood was collected by venipuncture. Serum and plasma were isolated via centrifugation of non-anticoagulated and heparinized blood, respectively, and stored at - 80C. For the current analyses, we included 128 participants who were enrolled between April and July, 2020 and who had at least one measurement on a binding assay or neutralization platform.

### Antibody assays

Supplementary Table 1 describes each of the assays performed as part of this analysis, including their sensitivity and specificity (as reported by the manufacturers for commercial assays, or by validation testing for non-commercial research assays). Commercially available platforms included the Abbott ARCHITECT (IgG), Ortho Clinical Diagnostics VITROS (IgG and total Ig), DiaSorin LIAISON (IgG), Roche Elecsys (total Ig), and Monogram PhenoSense (neutralizing antibodies) assays according to manufacturer specifications. Individuals living with HIV infection were excluded from analyses involving the Monogram PhenoSense assay, which utilizes an HIV backbone for the pseudovirus. Non-commercial research use assays included the Luciferase Immunoprecipitation Systems (LIPS) assay (total Ig) targeting the nucleocapsid (N) protein and the receptor binding domain (RBD) of the spike (S) protein performed in the Burbelo laboratory (additional raw data on responses to full S protein, highly correlated with RBD reponses, are included in Supplementary Tables 2 and 3), ^19^ the split luciferase assay (total Ig) targeting N and S performed in the Wells laboratory, ^20^ and the Luminex assay (IgG) targeting N (one full-length and one fragment), S, and RBD performed in the Greenhouse laboratory.

For the research use Luminex assay, we used a published protocol with modifications.^21^ Plasma samples were diluted to 1:100 in blocking buffer A (1xPBS, 0.05% Tween, 0.5% bovine serum albumin, 0.02% sodium azide). Antigens were produced using previously described constructs.^22,23^ Antigen concentrations used for COOH-bead coupling were as follows: S, 4 ug/mL; RBD, 2 ug/mL; and N, 3 ug/mL. Concentration values were calculated from the Luminex median fluorescent intensity (MFI) using a plate-specific standard curve consisting of serial dilutions of a pool of positive control samples. Any samples with MFIs above the linear range of the standard curve were serially diluted and rerun until values fell within range to obtain a relative concentration. A cutoff for positivity was established for each antigen above the maximum concentration value observed across 114 pre-pandemic SARS-CoV-2 negative control samples tested on the platform.

### Statistical Analyses

#### Comparing individuals across assays and estimating time to seroreversion

For each assay, we fit a linear mixed effects model that included a patient-specific random intercept. Given the longitudinal nature of our data set, we fit mixed effects models to explicitly account for the repeated measurement of individuals over time. We log-transformed the response variable for a subset of the assays based on assessment of their correlations with log-transformed neutralization titers (Supplementary Table 1). For observation *h* for individual *i*, we modeled their antibody response *Y*_*hi*_ on each assay as follows:

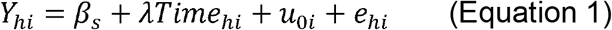

In Equation 1, β_*s*_ represents the overall mean for severity class *s*, where *s* was dichotomized into whether an individual was hospitalized or not-hospitalized; λ represents the fixed effect of *Time*_*hi*_, where *Time*_*hi*_ is data on the time since symptom onset (if symptomatic) or since positive PCR test (if asymptomatic). In addition, *u*_*0i*_ represents an individual-level random effect that is normally distributed with a mean of 0 and a standard deviation of τ, and *e*_*hi*_ represents the residual error that is normally distributed with a mean of 0 and a standard deviation of σ. We also considered models that included additional fixed effects for covariates such as age, ethnicity, sex, and HIV status (Supplementary Table 4). We did not find consistent differences in the slope of the antibody responses (λ) by hospitalization status across the majority of the assays used here; therefore we used a single slope for each assay throughout (Supplementary Table 5).

Since the timing of the baseline visit was variable between individuals, in order to directly compare the magnitude of measured responses for individuals on each assay, we used the mixed-effects model to estimate the antibody response that each person would have at 21 days post symptom onset (random intercept).^24^ We also used the model estimates to calculate the mean time to seroreversion *T* for severity class *s* on each assay, given the cutoff value for positivity (Supplementary Table 1), as follows:

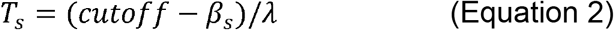

We performed bootstrapping to obtain 95% confidence intervals of *T*_*s*_ for each of the 14 assays. We used the time to seroreversion as the outcome here rather than alternative quantities such as the half-life, as the serologic responses obtained here did not all necessarily represent direct measurements of antibody titers. These models were fit using the *lme4* package using the R statistical software (https://www.R-project.org/).

#### Random forests modeling of demographic/clinical predictors and antibody responses

For each assay, we used random forests to model antibody responses based on 50 demographic and clinical predictors (Supplementary Table 6). We dichotomized the antibody response for each individual on each assay based on whether or not their estimated random intercept was in the top half or the bottom half of all fitted random intercepts on that assay. We first fit models to these dichotomized antibody responses using all available predictors; subsequently, we fit models to these dichotomized antibody responses on a down-selected set of predictors selected based on variable importance (i.e., mean decrease in accuracy). We quantified prediction accuracy using the out-of-bag error rate and the area under the curve (AUC). These models were built using the *randomForest* package using the R statistical software (version 3.5.3).

#### Estimating time-varying assay sensitivity

For each assay, we fit an extension of the linear mixed effects models described in Equation 1 above in a Bayesian hierarchical modeling framework, where we we allowed the standard deviation of the random intercept (τ) to be severity-specific (now referred to as τ_*s*_). For parsimony, we assumed the slopes, λ, to again be shared across all individuals as described above. To estimate changes in assay sensitivity over time, we simulated population distributions of Ŷ by iteratively sampling values from each of the posterior distributions of β_*s*_, λ, τ_*s*_, and σ. For each sampled value of τ_*s*_ and σ, we then sampled draws of *u*_*0i*_ and *e*_*hi*_ and determined the assay sensitivity at various time points (0, 60, 120, and 180 days after seroconversion) as the proportion of overall draws where Ŷ was above the cutoff value for positivity. We ran 4 Markov chain Monte Carlo (MCMC) chains of length 2,000 each using the Stan programming language (https://mc-stan.org/), and assessed convergence using the Gelman-Rubin Rhat statistic. We used uninformative priors for all parameters and hyper-parameters.

Data and code to reproduce all analyses are available at: https://github.com/EPPIcenter/liinc-Ab-dynamics.

### Ethical considerations

All participants signed a written informed consent form. The study was approved by the University of California, San Francisco Institutional Review Board.

## RESULTS

### Participant demographics and characteristics

As shown in Table 1, the cohort of 128 participants had an average age of 48 years (range 19-85 years), was relatively balanced in terms of sex (45% female at birth), and 26% of participants self-identified as being of Latinx ethnicity, a group which has been identified to be at-risk for COVID-19. Common medical comorbidities were hypertension (23%), lung disease (16%), and diabetes (13%). Notably, 18 individuals (14%) were living with HIV infection; 17 of 18 were on antiretroviral therapy. 121 individuals (95%) reported symptoms during their COVID-19 illness, but only 32 (25%) required hospitalization. Among those who had been hospitalized, 84% required supplemental oxygen, 42% required ICU admission, and 13% required mechanical ventilation. A minority of individuals (n=7, 5%) were asymptomatic.

**Table 1:** Demographic and clinical characteristics of the study participants.

| Characteristic | N=128 (%) |
| --- | --- |
| **Age (years)** | 47.8 (range: 19-85) |
| **Female sex at birth** | 57 (44.5) |
| **Race** |  |
| American Indian or Alaska Native | 4 (3.1) |
| Asian | 15 (11.7) |
| Black or African American | 7 (5.5) |
| Native Hawaiian or Other Pacific Islander | 3 (2.3) |
| White | 81 (63.4) |
| Declined | 20 (15.6) |
| **Latinx Ethnicity** | 33 (25.8) |
| **Medical Comorbidities** |  |
| Autoimmune disease | 9 (7.0) |
| Active cancer | 3 (2.3) |
| Diabetes | 17 (13.3) |
| HIV | 18 (14.1) |
| Heart disease | 3 (2.3) |
| Hypertension | 29 (22.7) |
| Lung disease | 21 (16.4) |
| Kidney disease | 2 (1.6) |
| Obesity (BMI ≥ 30) | 38 (29.7) |
| **Clinical Manifestations of COVID-19** |  |
| **Asymptomatic** | 7 (5.5) |
| **Symptomatic** | 121 (94.5) |
| Fever | 86 (70.5) |
| Chills | 75 (61.5) |
| Fatigue | 110 (90.2) |
| Cough | 89 (73.0) |
| Shortness of breath | 77 (63.1) |
| Rhinorrhea | 58 (47.5) |
| Sore throat | 56 (45.9) |
| Myalgias | 84 (68.9) |
| Nausea | 36 (29.5) |
| Vomiting | 12 (9.8) |
| Diarrhea | 50 (41.0) |
| Anosmia or dysgeusia | 82 (67.2) |
| Headache | 77 (63.1) |
| **Hospitalized** | 31 (24.2) |
| Required supplemental oxygen | 26 (83.9) |
| Required ICU admission | 13 (41.9) |
| Required mechanical ventilation | 4 (12.9) |
| **Numbers of symptoms reported** |  |
| 1 to 3 | 13 (10.2%) |
| 4 to 6 | 34 (26.6%) |
| 7 to 9 | 47 (36.7%) |
| 10 to 13 | 27 (21.1%) |
| **Enrollment and follow-up** |  |
| Baseline visit, days since onset (median) | 63 (range: 22-157) |
| Follow-up time, days since onset (median) | 110 (range: 22-157) |
| Time points contributed (median) | 2 (range: 1-4) |

The baseline visit for participants occurred at a median of 63 days (range: 22 - 157) after symptom onset (Supplementary Figure 1). Participants contributed a median of 2 samples each (range: 1 - 4) and were followed-up for a median of 110 days after symptom onset (range: 22 - 157). A total of 267 samples from the 128 enrolled participants were tested using at least one of the 14 assays evaluated; 171 samples from 88 individuals were tested using all 14 assays (Supplementary Table 1, Supplementary Figure 2). Individuals with HIV were excluded from the Neut-Monogram as noted above.

### Substantial heterogeneity in antibody responses across individuals and assays

We observed substantial heterogeneity in measured antibody responses in individuals at baseline and throughout follow-up across all assays (Figure 1, raw data available in Supplementary Tables 2 and 3). We observed variable trajectories of antibody responses between assays, with some (N-Abbott, N-Split Luc, S-Ortho IgG, and Neut-Monogram) showing a clear decrease over time, other assays (S-Ortho Ig and N-Roche) showing a clear increase, and the remainder with more stable values (Figure 1, Supplementary Table 7). When comparing antibody levels between individuals, responses were very heterogeneous, with some individuals mounting strong responses for all assays and others with weak responses even at the initial visit (below the positivity cutoff for some assays).

**Figure 1:**
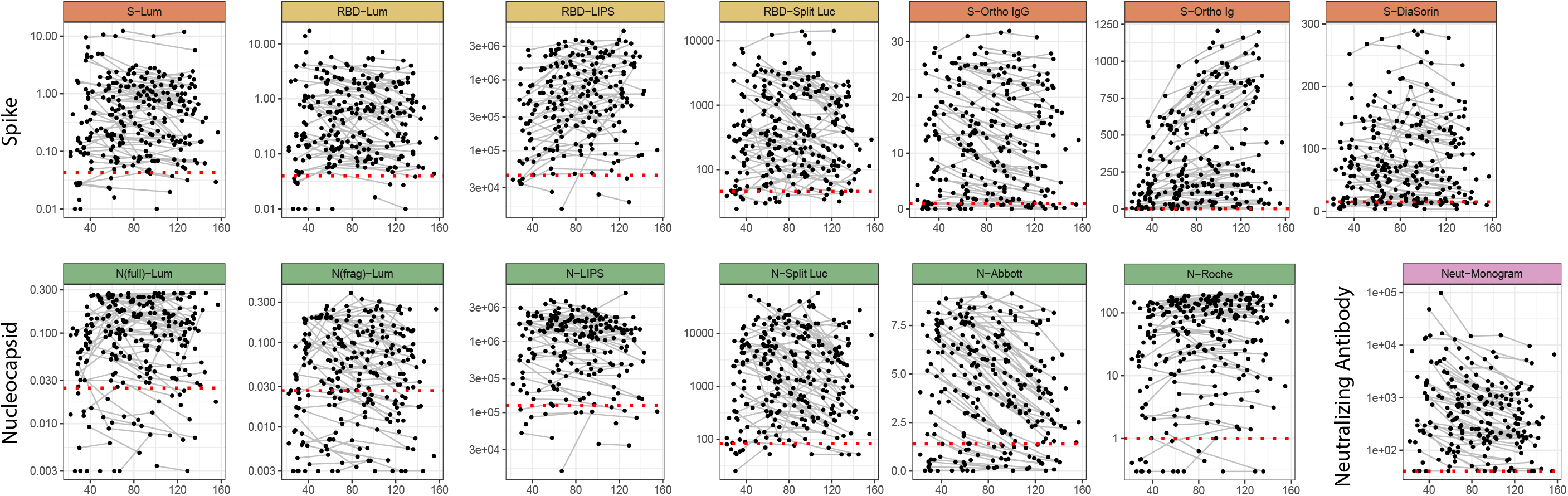
Longitudinal antibody kinetics. Time since symptom onset is shown on the x-axis versus the measured antibody response for each assay. For asymptomatic individuals, the time since the first positive PCR test was used. Black points indicate individual time points, and longitudinal samples are connected with grey lines. Y-axes are transformed as indicated in Supplementary Table 1. Red dotted lines indicate cutoff values for positivity, as indicated in Supplementary Table 1.

### Strong correlation between binding and neutralization assays

We observed high levels of correlation between estimated antibody levels at 21 days post symptom onset (random intercept) for all assays, with Spearman correlations ranging between 0.55 and 0.96 (Figure 2A, Supplementary Figure 3). Rank correlations were consistently higher between binding assays using the same antigenic target (S/RBD vs. N) than between those using different targets, despite the variety in platforms used and the measurement of responses to both targets on some platforms (LIPS, Luminex, split luciferase). Titers of neutralizing antibodies correlated well with all binding assays (range: 0.60 to 0.88) and correlated most highly with responses to the S protein (range: 0.76 to 0.88), as might be expected given the expression of S protein on the pseudovirus used in the neutralization assay (Figure 2B, Supplementary Figure 3). We found no substantive differences in correlations between binding and neutralization assays at timepoints before vs. after 90 days, suggesting these relationships did not appreciably change over the duration of observed follow-up (Supplementary Table 8).

**Figure 2:**
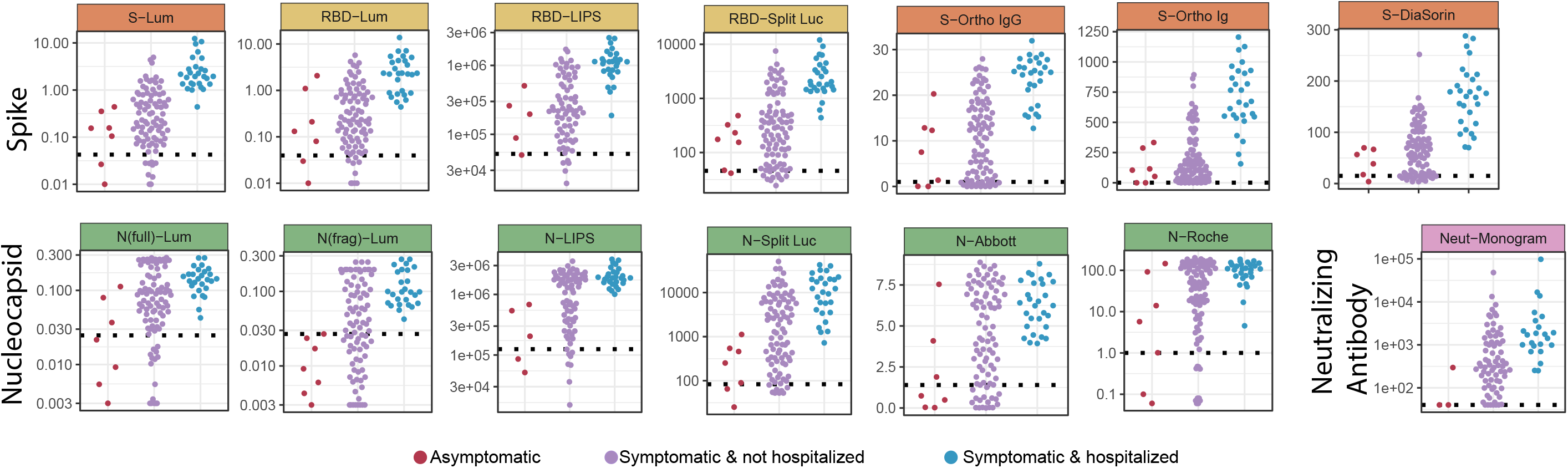
Correlation of responses between assays. **(A)** Spearman correlation of random intercepts derived from a mixed effects model, representing responses at day 21 for each individual from the longitudinal data. Assays are sorted by hierarchical clustering using average distance clustering. Darker blue indicates higher correlation; colored label box indicates antigen for each binding assay and the neutralizing assay. **(B)** Pairwise scatterplots showing the random intercepts for the neutralizing assay (x-axis) versus the random intercepts for each of the other assays (y-axis).

### Disease severity is strongly associated with the magnitude of antibody responses

Baseline antibody responses for each study participant showed remarkably consistent patterns across all assays when stratified by severity class, with asymptomatic individuals having the lowest responses, hospitalized individuals having the highest, and symptomatic but not hospitalized individuals having intermediate responses (Figure 3). While the number of asymptomatic individuals was small, responses were significantly lower in these individuals than those who were symptomatic but not hospitalized for multiple assays; hospitalized individuals had significantly higher responses than both other groups for all assays with the notable exception of the neutralization assay (Supplementary Table 9). Despite these consistent patterns, there was still substantial variation in the magnitude of responses between participants within each severity category. Notably, age, sex, HIV status, and Latinx ethnicity showed little association with antibody responses after adjusting the analysis for hospitalization (Supplementary Table 4).

**Figure 3:**
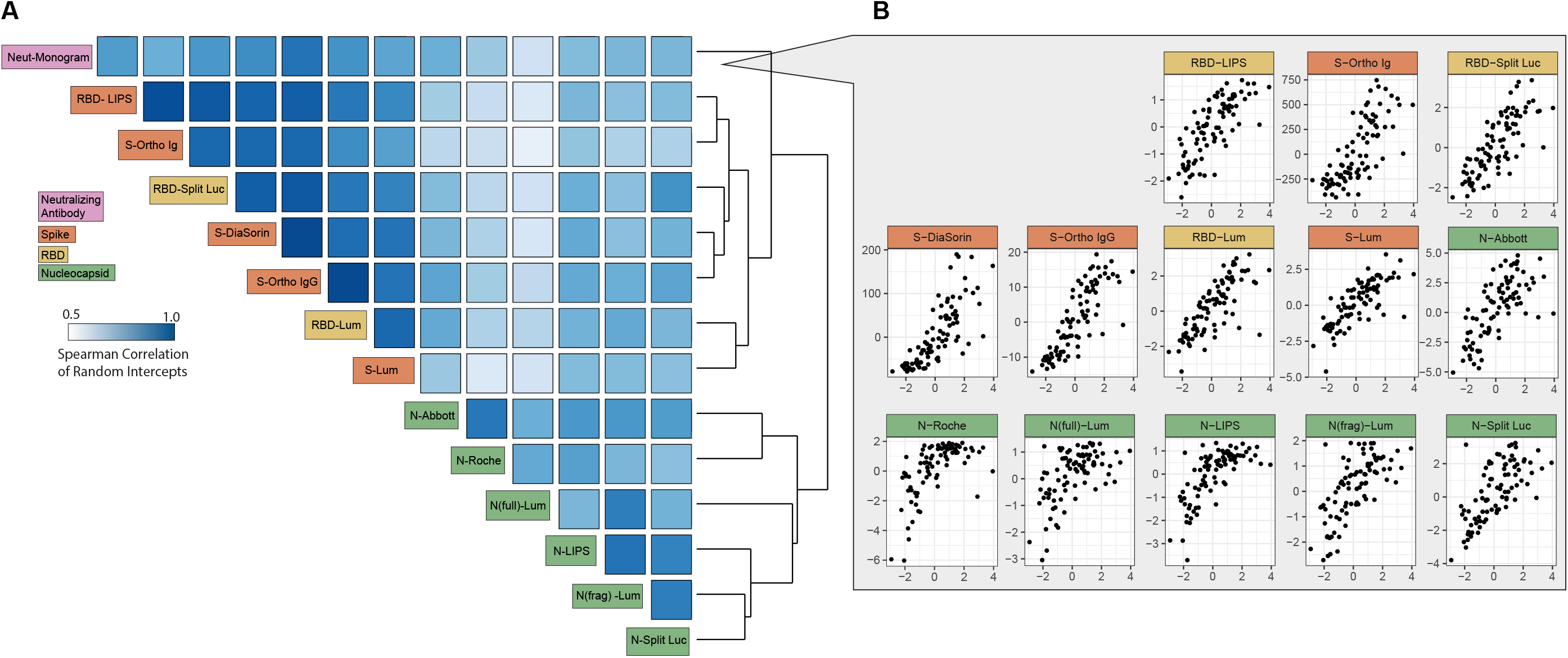
Severity-stratified antibody response at baseline. Swarm plot of antibody response at the baseline visit for each study participant by assay, stratified into individuals who experienced no symptoms, individuals who experienced symptoms but were not hospitalized, and those who experienced symptoms and were hospitalized. Y-axes are transformed as indicated in Supplementary Table 1.

### Need for hospitalization, cough, and fever are key predictors of antibody responses

We next examined which of 50 individual demographic and clinical variables were the strongest predictors of the magnitude of the antibody response (top vs. bottom half of responders for each assay, Supplementary Figure 4) using a random forests algorithm. Among the entire cohort (n=128), the presence and duration of cough and fever, and need for hospitalization and supplemental oxygen during the initial illness, were the most important predictors of the antibody response (Figure 4A). The ranks of their importance varied subtly but were largely consistent across the 14 assays evaluated, and random forests models including only these 6 variables were able to predict high versus low magnitude of response on each assay with reasonably high accuracy (AUCs ranging from 0.74 to 0.86, Supplementary Table 10). Among those individuals who were not hospitalized (n=96), presence and duration of fever and cough remained the most important predictors of a high antibody response (Figure 4B). These four variables alone were predictive of high vs. low responses within this subset of individuals with modest accuracy (all except RBD-LIPS with AUCs above 0.6).

**Figure 4:**
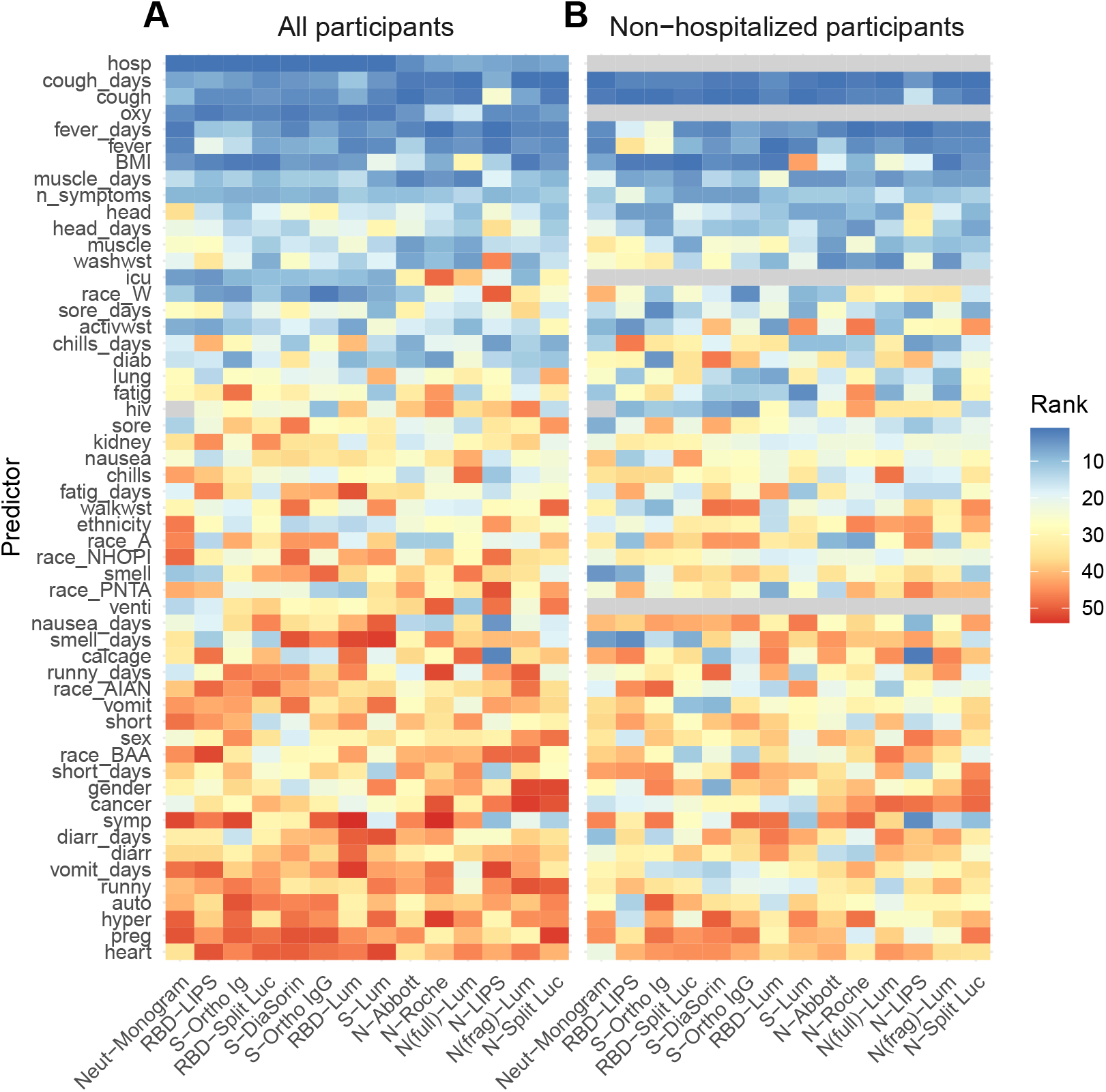
Clinical predictors of antibody responses. Rank of variable importance (1=highest rank, 50=lowest rank) in a random forest classifier of top half vs. bottom half of responders for each assay, based on random intercepts, including **(A)** all individuals (n=128) and **(B)** only individuals who were not hospitalized (n=96), as hospitalization is a strong predictor of antibody response. Variable importance was determined as the reduction in classification error averaged across 10 runs of the algorithm. Variables only relevant to hospitalized individuals (i.e., whether or not the individual was hospitalized, whether or not oxygen was required, whether or not the individual was in the ICU, and whether or not the individual required a ventilator) were omitted from the classifier in panel B and shown in grey. In addition, HIV status is excluded as a predictor for the Neut-Monogram assay for reasons described in the main text. The dependent variable (individual-level random intercepts derived from a mixed effects model) is dichotomized into “high” and “low”, determined by the random intercept being in the upper or lower half of all random intercepts for that assay, respectively. Full labels of the predictor variables are provided in Supplementary Table 6.

### Time to seroreversion varies considerably across platforms and by infection severity

Using the mixed effect model described above, we estimated the expected time to seroreversion (when antibodies would become undetectable for an average individual) for each platform, assuming that antibody responses changed linearly over time. We estimated separate times to seroreversion for hospitalized and non-hospitalized individuals, since hospitalization status was a strong predictor of baseline antibody status. Estimated time to seroreversion was substantially shorter for non-hospitalized vs. hospitalized individuals for all assays, consistent with the lower initial antibody titers in those individuals. For those assays where antibody levels decreased over time, we also observed marked variation in times to seroreversion between assays, ranging from 96 days for N(frag)-Lum to 925 days for S-DiaSorin; the estimated time to seroreversion is infinity for RBD-LIPS, S-Ortho Ig, and N-Roche, which exhibited increasing mean antibody responses over time in this dataset (Figure 5A, Supplementary Table 7).

**Figure 5:**
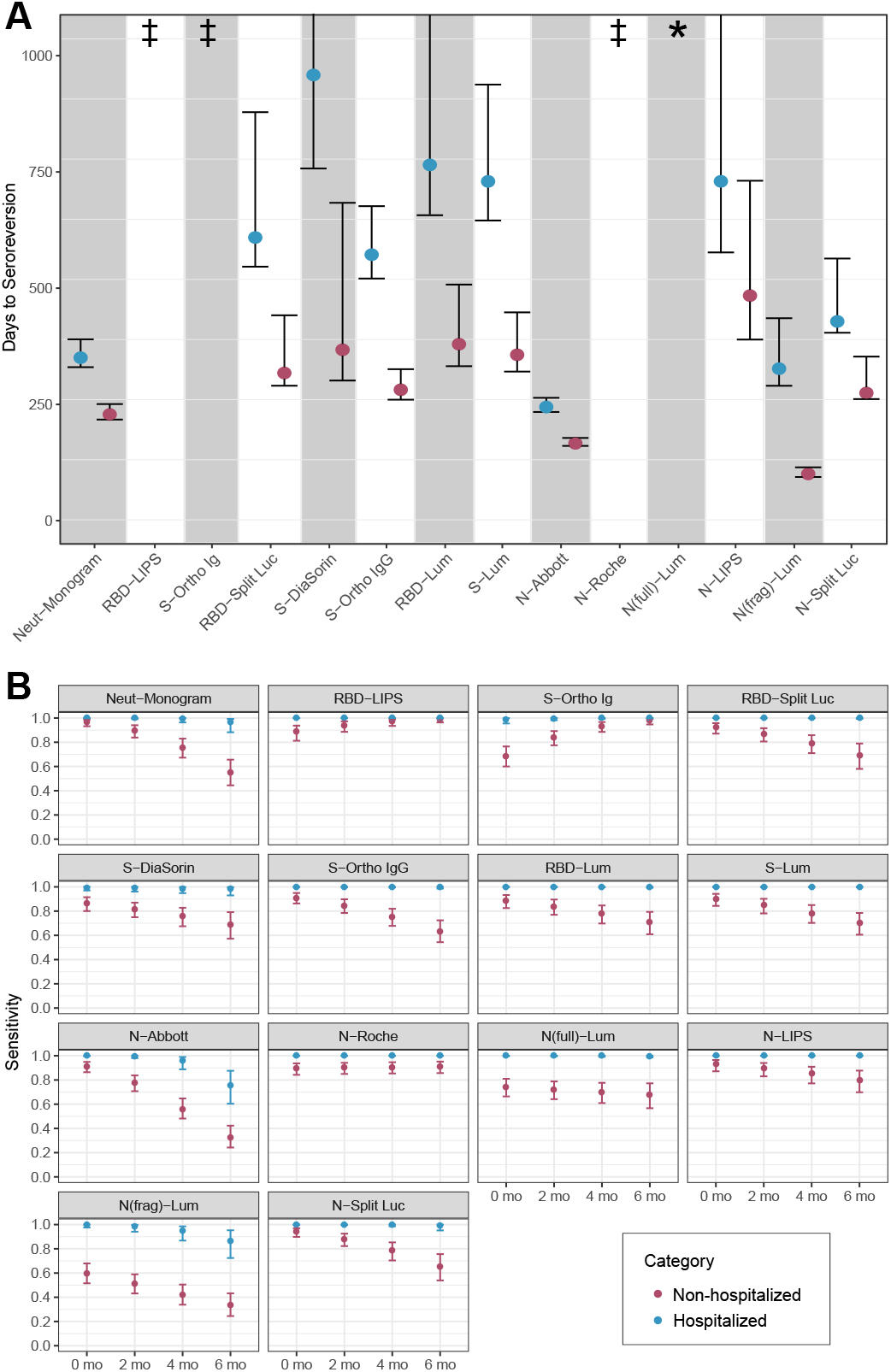
Estimated time to seroreversion and assay sensitivity by time and hospitalization status. **(A)** Mean time to seroreversion for individuals tested on each assay, stratified by hospitalization status, with 95% confidence intervals derived from bootstrapping. Symbol (‡) indicates increasing antibody responses over time (95% confidence interval for time to seroreversion was negative and did not cross 0), and symbol (*) indicates antibody responses for which 95% confidence interval of time to seroreversion crossed 0. **(B)** Estimated sensitivity of each assay (showing posterior median estimates and 95% credible intervals), stratified by hospitalization status at 2 month intervals, from 0 months to 6 months after seroconversion. Seroconversion was assumed to occur (if at all) 21 days after symptom onset (if symptomatic) or 21 days after positive PCR test (if asymptomatic).

### Sensitivity of assays to detect prior infection varies as a function of time and infection severity

We next assessed how sensitivity of each platform varied as a function of time and disease severity. Because the sample size of asymptomatic individuals was small, they were grouped with other non-hospitalized individuals for this analysis (Supplementary Figure 5). We found considerable heterogeneity in sensitivity between assays and as a function of illness severity and time since infection. Across all fourteen assays, sensitivity at each time point was higher in the hospitalized subset of the cohort than in the non-hospitalized subset, the latter group representing the majority of infections in the general population (Figure 5B, Supplementary Table 11). The magnitude of this difference varied between assays and also over time, and was often considerable (Supplementary Figure 6). Estimated sensitivity declined over time for eleven of the fourteen assays, but increased for RBD-LIPS, S-Ortho Ig, and N-Roche, consistent with the observed increase in magnitude of response over time for these assays. Overall, RBD-LIPS showed the most consistently high sensitivity over time and the smallest difference between hospitalized and non-hospitalized individuals, ranging from 88% (95% credible interval: 81% - 94%) at month 0 to 99% (95% credible interval: 96% - 100%) at month 6 in non-hospitalized individuals. Of the remaining research-use assays, N-LIPS, RBD- and N-Split Luc, and RBD- and S-Lum showed a similar pattern, with high sensitivity initially followed by a decline in sensitivity amongst non-hospitalized individuals over time; N(full)-Lum and N(frag)-Lum had consistently poor sensitivity for non-hospitalized individuals. All commercial assays performed similarly well during early convalescence, albeit with lower sensitivity for non-hospitalized individuals for S-DiaSorin and S-Ortho Ig. N-Abbott showed the greatest decline in sensitivity with time and the greatest difference between hospitalized and non-hospitalized participants, with sensitivity varying from 100% (95% credible interval: 100% - 100%) in hospitalized individuals soon after infection to 33% (95% credible interval: 24% - 42%) in non-hospitalized individuals at 6 months. Of note, neutralization titers remained detectable for nearly all hospitalized individuals up to six months, but were estimated to become undetectable on this assay for nearly half of non-hospitalized individuals by this time point.

### Varying assay sensitivity affects interpretation of individual antibody test results

Lastly, negative predictive values were calculated for the commercial assays to illustrate potential effects of these changes in sensitivity if the assays were used to ascertain prior infection in individuals (assuming values of specificity as reported by the manufacturers). As expected, negative predictive values decreased with increasing prevalence and, except for S-Ortho Ig and N-Roche, time (Supplementary Figure 7).

## DISCUSSION

This study documents the large heterogeneity in longitudinal antibody responses to SARS-CoV-2 across a large number of commercial and research assays in a diverse cohort of individuals. Measured responses in all binding assays correlated well with each other and, particularly for those measuring responses to spike protein, with pseudovirus neutralization. For all assays, we found a consistent, strong, and dose-dependent effect of disease severity on antibody magnitude. Despite these similarities, assays performed quite differently in terms of sensitivity to detect prior infection and in the durability of measured responses, leading to large discrepancies in sensitivity between assays in the months following infection. Thus, the ability to detect previous infection by SARS-CoV-2 using an antibody test is highly dependent on the severity of the initial infection, when the sample is obtained relative to infection, and the assay used.

Prior work has shown that antibody responses in individuals with symptomatic COVID-19 have in some cases been associated with disease severity.^4–12^ We observed significant variability in antibody responses between study participants which was largely explained by the self-reported symptom constellation and the severity of the acute illness. A few simple variables consistently predicted the magnitude of the antibody responses across multiple assay platforms and antigen targets; these symptoms (e.g., fever, cough) are similar to those recently described in a population-based Icelandic cohort.^6^ Importantly, in contrast to that cohort, characteristics like age and sex were not predictive of these responses, after accounting for disease severity.

We also observed substantial heterogeneity between assays in terms of overall sensitivity, particularly over time. These findings build upon prior work showing differences in sensitivity during early convalescence.^7,10^ This finding may also provide insight into apparent discrepancies in previous studies that have reported different durabilities of antibody responses.^25,26^ Differences in performance were particularly pronounced amongst non-hospitalized individuals, who have lower antibody responses and comprise the majority of those infected with SARS-CoV-2. Notably, antibody responses in such individuals are expected to be reliably detectable over 6 months in only 2 of the commercial binding assays tested – S-Ortho Ig and N-Roche – which were the only to employ direct detection format of antibodies (to different viral targets). Interestingly, these 2 assays, along with a research-use assay employing direct detection (RBD-LIPS), were the only ones to demonstrate increasing rather than decreasing antibody signal over time, possibly due to continued affinity maturation of the antibody response^27^ playing a larger role in detection with this format. This is in contrast to the indirect format assays and neutralization assay, all of which demonstrated waning over time. Another possible explanation for discrepancies between assays is that only assays with the highest signal to noise ratio are able to consistently detect antibodies above background in those with the lowest titers, i.e. those multiple months out from mild infections. This variation in sensitivity is relevant for several reasons. First, it provides further evidence that use-cases need to be considered when evaluating performance of antibody tests. While all evaluated assays had near perfect sensitivity for detecting antibody responses among hospitalized individuals and therefore could be useful as an indirect diagnostic tool in that setting, their sensitivity to detect responses in the general population, where most infections are mild, is much lower and quite variable.^1^ Second, it implies that using assays with decreasing sensitivity over time for population seroprevalence studies will underestimate the true proportion of previously infected individuals, and that this underestimation will be more substantial as the amount of time that has passed since infection increases. Third, it shows dramatic differences between the sensitivity reported by test manufacturers, often limited to validation sample sets from hospitalized and/or recently infected individuals that were readily available early in the pandemic, and the expected sensitivity in the general population. Because of this, our study provides information that could be useful for assay selection when planning future serosurveys, and could help correct the interpretation of large-scale population-based seroprevalence studies that have used some of these assays. Finally, individual patients or providers utilizing these assays to assess the presence or absence of prior infection and/or immune status should take these considerations into account, given the poor negative predictive value of some tests.

This analysis has several notable strengths, including the utilization of a broad array of antibody platforms at multiple time points in a diverse cohort of individuals with varying degrees of illness severity and rich clinical phenotyping. However, there are several limitations. The cohort was not population-based and therefore not truly representative of all individuals with SARS-CoV-2 infection. Despite this, we endeavored to recruit a cohort that spanned the spectrum of SARS-CoV-2 infection. Second, this analysis included only a small number of asymptomatic individuals, which are likely to differ from symptomatic patients in terms of initial and possibly longitudinal responses based on our limited data and prior results;^11^ additional asymptomatic individuals are being recruited for future analyses. Third, the current analysis is limited to samples obtained up to four months after infection. Data from longer follow-up times will be very useful in order to estimate the longer term kinetics of antibody responses in all platforms with more certainty. The assumption of linearity, while empirically appropriate on the time-scales of data that we have here, may not be accurate for extrapolation into the distant future as antibody responses often follow more complex dynamics of boosting and waning over time.^28^ Finally, it is important to recognize that assays optimally suited for serosurveillance may not be equally suitable for other use-cases, such as identifying recent infection, detecting reinfection, determining protective capacity, or determining potency of COVID-19 convalescent plasma.^29^ Evaluating the performance of assays for each of these use-cases will require different study designs and sample sets.

As SARS-CoV-2 vaccination becomes a reality, many serosurveillance efforts will need to increasingly rely upon assays that can distinguish vaccination from natural infection, especially when it is not possible to obtain additional information on the vaccination status of participants. Currently distributed vaccines in the US are based on S protein, thus responses to S-based assays will likely reflect a combination of natural infection and vaccination, whereas assays based on N will reflect only natural infection. While several of the N-based assays we evaluated performed well, one of the two commercially available N-based assays demonstrated substantial waning of sensitivity over time which will affect estimates of the seroprevalence of natural infection, but could be a more useful indicator of recent infection along with other potential markers such as IgM. Further studies will be needed in order to characterize the performance of these and other assays for serosurveillance in the presence of vaccine induced immunity.

## Conclusions

In this study, we demonstrated substantial differences in the detectability of antibody responses to SARS-CoV-2 related to illness severity, time since infection, and assay platform. These results will be important in choosing and interpreting serologic assays for evaluating infection and immunity in population surveillance studies.

## Data Availability

Raw antibody data at the participant and visit level are provided in Supplementary Tables 2 and 3.

## ACKNOWLEDGEMENTS

We are grateful to the LIINC study participants and to the clinical staff who provided care to these individuals during their acute illness period. We acknowledge LIINC study team members Tamara Abualhsan, Mireya Arreguin, Jennifer Bautista, Monika Deswal, Heather Hartig, Marian Kerbleski, Lynn Ngo, Fatima Ticas, and Meghann Williams for their contributions to the program. We are grateful to Khamal Anglin, Grace Bronstone, Jessica Chen, Michelle Davidson, Kevin Donohue, Peyton Ellis, Sarah Goldberg, Scott Lu, Jonathan Massachi, Sujata Mathur, Irum Mehdi, Victoria Wong Murray, Enrique Martinez Ortiz, Jesus Pineda-Ramirez, Mariela Romero, Paulina Rugart, Hannah Sans, Joshua Shak, Jaqueline Tavs, and Jacob Weiss for assistance with data entry and validation. We thank Ms. Heather Tanner (Vitalant Research Institute) for technical assistance.

## DATA AVAILABILITY

Raw antibody data at the participant and visit level are provided in Supplementary Tables 2 and 3.

## AUTHOR CONTRIBUTIONS

MJP, JDK, RLR, NJM, SGD, TJH, IRB, and BG oversee the LIINC cohort, which is funded by MAS, MG, SGD, and TJH. MJP, ST, JH, TJH, IRB, and BG designed the study and analysis plan. MJP, LT, RH, VT, EF, and YH collected clinical data using study instruments designed by MJP, JDK, RH, SGD, and JNM. LT, NSI, KT, OJ, SEM, CT, CCN processed and stored biological specimens. LT, NSI, KT, OJ, SEM, CT, CCN, M-AP, AV, MAR, JP, JH, LT, TW, CJP, CYC, PJN, CDG, MS, MPB, SKE, XXZ, JAW, AS, TWK, JEP, WW, PDB, JIC, TJH, IRB, and BG performed or oversaw performance of antibody measurements in their respective laboratories. MJP, ST, JH, TJH, IRB, and BG analyzed and interpreted the raw data and drafted the manuscript. JDK, M-AP, AV, MAR, JP, JH, TW, CJP, CYC, PJN, CD, MS, MPB, SKE, JAW, TWK, PDB, RLR, JNM, SGD provided substantial scientific critiques and recommendations regarding the interpretation of the data. All authors edited and approved the final version of the manuscript.

## SUPPLEMENTARY INFORMATION

**Supplementary Table 1: Description of each assay**. Unit abbreviations: S/C = sample result to calibrator result index; COI = cutoff index; AU/mL = arbitrary unit per mL; ID50 = 50% inhibitory dilution; RLU = relative light unit; LU = light unit; conc = relative concentration. Symbol (*) indicates that the cutpoint for Neut-Monogram is the lower limit of detection for the assay. Antigen abbreviations: N = nucleocapsid; S = spike; RBD = receptor binding domain.

**Supplementary Table 2: Raw data at the patient level**. Patient ID, severity class, binned age in years, and sex.

**Supplementary Table 3: Raw data at the sample level**. Patient ID, time since symptom onset (or for asymptomatic individuals, time since the first positive PCR test), and antibody response for each of the 15 assays (including S-LIPS, which was highly correlated with RBD-LIPS).

**Supplementary Table 4: Outputs of regression models evaluating the association between demographic variables and antibody levels, controlling and not controlling for hospitalization**. Severity is characterized by hospitalization status (reference group: hospitalized). Demographic covariates considered for inclusion as population-level intercepts are HIV status (reference group: HIV+), sex (reference group: male), ethnicity (reference group: Hispanic), and age (reference group: older than or equal to 44). Log transformations of the data performed if indicated in Supplementary Table 1.

**Supplementary Table 5: Outputs of models testing for interaction between hospitalization status and slope in the linear mixed effects models**. Columns indicate the assay and estimate of the additional contribution of being hospitalized to the slope (Δλ_hosp_), with 95% confidence intervals and p-values.

**Supplementary Table 6: Clinical variable shorthands and corresponding REDCap questions**. Variable name, question asked, and categories (if applicable).

**Supplementary Table 7: Outputs of the linear mixed effects models by assay**. Point estimates of the parameters from the linear mixed effects models: population-level slope (λ), population-level intercept for hospitalization status *s* (β_s_), days to seroreversion (*T*_s_) stratified by hospitalization status, τ, and σ. Log transformations of the data performed if indicated in Supplementary Table 1.

**Supplementary Table 8: Correlations of binding assays with pseudotype virus neutralization**. Spearman rank correlation coefficients and 95% confidence intervals derived from bootstrapping, comparing (A) random intercepts, (B) all raw data, (C) all raw data from less than 90 days after symptom onset (‘early’), and (D) all raw data from greater than 90 days after symptom onset (‘late’).

**Supplementary Table 9: Outputs of pairwise t-tests of antibody responses at baseline between severity groups**. P-values < 0.05 depicted in bold.

**Supplementary Table 10: Classification accuracy of the random forest model when including a down-selected set of predictors**. The outcome for each individual is dichotomized as being in the top half vs. bottom half of random intercepts by assay. Variables included in the down-selected set for the model including all patients include: hospitalization status (binary), needing oxygen (binary), fever (binary), duration of fever (continuous), cough (binary), and duration of cough (continuous). Variables included in the down-selected set for the model including only non-hospitalized patients include: fever (binary), duration of fever (continuous), cough (binary), and duration of cough (continuous). The area under the curve (AUC) estimates are averaged across 10 runs of the algorithm.

**Supplementary Table 11: Assay sensitivity by time, severity, and assay**. Columns include the assay, hospitalization status, time, and assay sensitivity (posterior mean, standard error of the posterior mean, 2.5th quantile of the posterior distribution, posterior median, and 97.5th quantile of the posterior distribution).

**Supplementary Figure 1: Distribution of time since infection at baseline visit, by severity**. Days since symptom onset (if symptomatic) or positive PCR test (if asymptomatic) of the baseline visit, stratified by severity.

**Supplementary Figure 2: Sample inclusion for each platform. (A)** Of the 128 total individuals included, how many individuals had at least one sample tested on each assay. **(B)** Of the 267 total samples tested, how many samples were tested on each assay.

**Supplementary Figure 3: Correlation between all assays**. The values of the **(A)** random intercepts, **(B)** all raw data, **(C)** all raw data from less than 90 days after symptom onset (‘early’), and **(D)** all raw data from greater than 90 days after symptom onset (‘late’) are shown on the lower triangle. The Spearman rank correlation coefficients and p-values of the observed coefficients are shown on the upper triangle.

**Supplementary Figure 4: Distribution of the random intercept of antibody values for each individual by assay**. X-axis is the raw value of the random intercept from the linear mixed effects model. Red signifies random intercept values in the bottom half of that assay, and blue signifies random intercept values in the top half of that assay. These binary values were used as outcome variables in the random forests modeling.

**Supplementary Figure 5: Raw data by time, hospitalization status, and assay**. Raw antibody response data are either log-transformed or not transformed, according to Supplementary Table 1. The x-axis represents time since seroconversion in days, where seroconversion was assumed to occur (if at all) 21 days after symptom onset (if symptomatic) or 21 days after positive PCR test (if asymptomatic). The cutoff for positivity on that assay is shown by the dashed black line.

**Supplementary Figure 6: Ratio of sensitivity in non-hospitalized individuals to hospitalized individuals over time**. Posterior median estimates and 95% credible intervals shown.

**Supplementary Figure 7: Negative predictive values of the commercial assays**. Negative predictive values shown are based on the estimated assay sensitivities for non-hospitalized individuals in Figure 5B, for a range of prevalence between 5% and 50% (x-axis). Lower panels show the same data with a smaller range in the y axis to visualize small differences.

## REFERENCES

1. Takahashi, S., Greenhouse, B. & Rodríguez-Barraquer, I. Are Seroprevalence Estimates for Severe Acute Respiratory Syndrome Coronavirus 2 Biased? J. Infect. Dis. 222, 1772–1775 (2020).

2. Guan, W.-J. et al.Clinical Characteristics of Coronavirus Disease 2019 in China. N. Engl. J. Med. 382, 1708–1720 (2020).

3. Richardson, S. et al.Presenting Characteristics, Comorbidities, and Outcomes Among 5700 Patients Hospitalized With COVID-19 in the New York City Area. JAMA 323, 2052–2059 (2020).

4. Seow, J. et al.Longitudinal observation and decline of neutralizing antibody responses in the three months following SARS-CoV-2 infection in humans. Nat Microbiol 5, 1598–1607 (2020).

5. Long, Q.-X. et al.Clinical and immunological assessment of asymptomatic SARS-CoV-2 infections. Nat. Med. 26, 1200–1204 (2020).

6. Gudbjartsson, D. F. et al.Humoral Immune Response to SARS-CoV-2 in Iceland. N. Engl. J. Med. 383, 1724–1734 (2020).

7. Naaber, P. et al.Evaluation of SARS-CoV-2 IgG antibody response in PCR positive patients: Comparison of nine tests in relation to clinical data. PLoS One 15, e0237548 (2020).

8. Chen, Y. et al.Quick COVID-19 Healers Sustain Anti-SARS-CoV-2 Antibody Production. Cell 183, 1496–1507.e16 (2020).

9. Chen, X. et al.Disease severity dictates SARS-CoV-2-specific neutralizing antibody responses in COVID-19. Signal Transduct Target Ther 5, 180 (2020).

10. Kowitdamrong, E. et al.Antibody responses to SARS-CoV-2 in patients with differing severities of coronavirus disease 2019. PLoS One 15, e0240502 (2020).

11. Lei, Q. et al.Antibody dynamics to SARS-CoV-2 in asymptomatic COVID-19 infections. Allergy (2020) doi:10.1111/all.14622.

12. Zhao, J. et al.Antibody Responses to SARS-CoV-2 in Patients With Novel Coronavirus Disease 2019. Clin. Infect. Dis. 71, 2027–2034 (2020).

13. Ibarrondo, F. J. et al.Rapid Decay of Anti-SARS-CoV-2 Antibodies in Persons with Mild Covid-19. N. Engl. J. Med. 383, 1085–1087 (2020).

14. Muecksch, F. et al.Longitudinal analysis of clinical serology assay performance and neutralising antibody levels in COVID19 convalescents. medRxiv (2020) doi:10.1101/2020.08.05.20169128.

15. Ainsworth, M. et al.Performance characteristics of five immunoassays for SARS-CoV-2: a head-to-head benchmark comparison. Lancet Infect. Dis. (2020) doi:10.1016/S1473-3099(20)30634-4.

16. Lau, E. H. Y. et al.Neutralizing antibody titres in SARS-CoV-2 infections. Nat. Commun. 12, 63 (2021).

17. Kroenke, K., Spitzer, R. L. & Williams, J. B. W. The PHQ-15: validity of a new measure for evaluating the severity of somatic symptoms. Psychosom. Med. 64, 258–266 (2002).

18. Herdman, M. et al.Development and preliminary testing of the new five-level version of EQ-5D (EQ-5D-5L). Qual. Life Res. 20, 1727–1736 (2011).

19. Burbelo, P. D. et al.Sensitivity in Detection of Antibodies to Nucleocapsid and Spike Proteins of Severe Acute Respiratory Syndrome Coronavirus 2 in Patients With Coronavirus Disease 2019. J. Infect. Dis. 222, 206–213 (2020).

20. Elledge, S. K. et al.Engineering luminescent biosensors for point-of-care SARS-CoV-2 antibody detection. medRxiv (2020) doi:10.1101/2020.08.17.20176925.

21. Wu, L. et al.Optimisation and standardisation of a multiplex immunoassay of diverse Plasmodium falciparum antigens to assess changes in malaria transmission using sero-epidemiology. Wellcome Open Res 4, 26 (2019).

22. Amanat, F. et al.A serological assay to detect SARS-CoV-2 seroconversion in humans. Nat. Med. 26, 1033–1036 (2020).

23. Pilarowski, G. et al.Performance Characteristics of a Rapid Severe Acute Respiratory Syndrome Coronavirus 2 Antigen Detection Assay at a Public Plaza Testing Site in San Francisco. J. Infect. Dis. (2021) doi:10.1093/infdis/jiaa802.

24. Long, Q.-X. et al.Antibody responses to SARS-CoV-2 in patients with COVID-19. Nat. Med. 26, 845–848 (2020).

25. Rapid Decay of Anti-SARS-CoV-2 Antibodies in Persons with Mild Covid-19. N. Engl. J. Med. 383, e74 (2020).

26. Iyer, A. S. et al.Persistence and decay of human antibody responses to the receptor binding domain of SARS-CoV-2 spike protein in COVID-19 patients. Sci Immunol 5, (2020).

27. Gaebler, C. et al.Evolution of antibody immunity to SARS-CoV-2. Nature (2021) doi:10.1038/s41586-021-03207-w.

28. Huang, A. T. et al.A systematic review of antibody mediated immunity to coronaviruses: kinetics, correlates of protection, and association with severity. Nat. Commun. 11, 4704 (2020).

29. Denise M. Hinton, Chief Scientist, Food and Drug Administration, Office of the Assistant Secretary for Preparedness and Response, Office of the Secretary, U.S. Department of Health and Human Services. Letter of Authorization, Reissuance of Convalescent Plasma EUA February 4, 2021. (2021).

