## Supplementary figures and images for "SARS-CoV-2 antibody magnitude and detectability are driven by disease severity, timing, and assay"

### Supplemental Figure 1

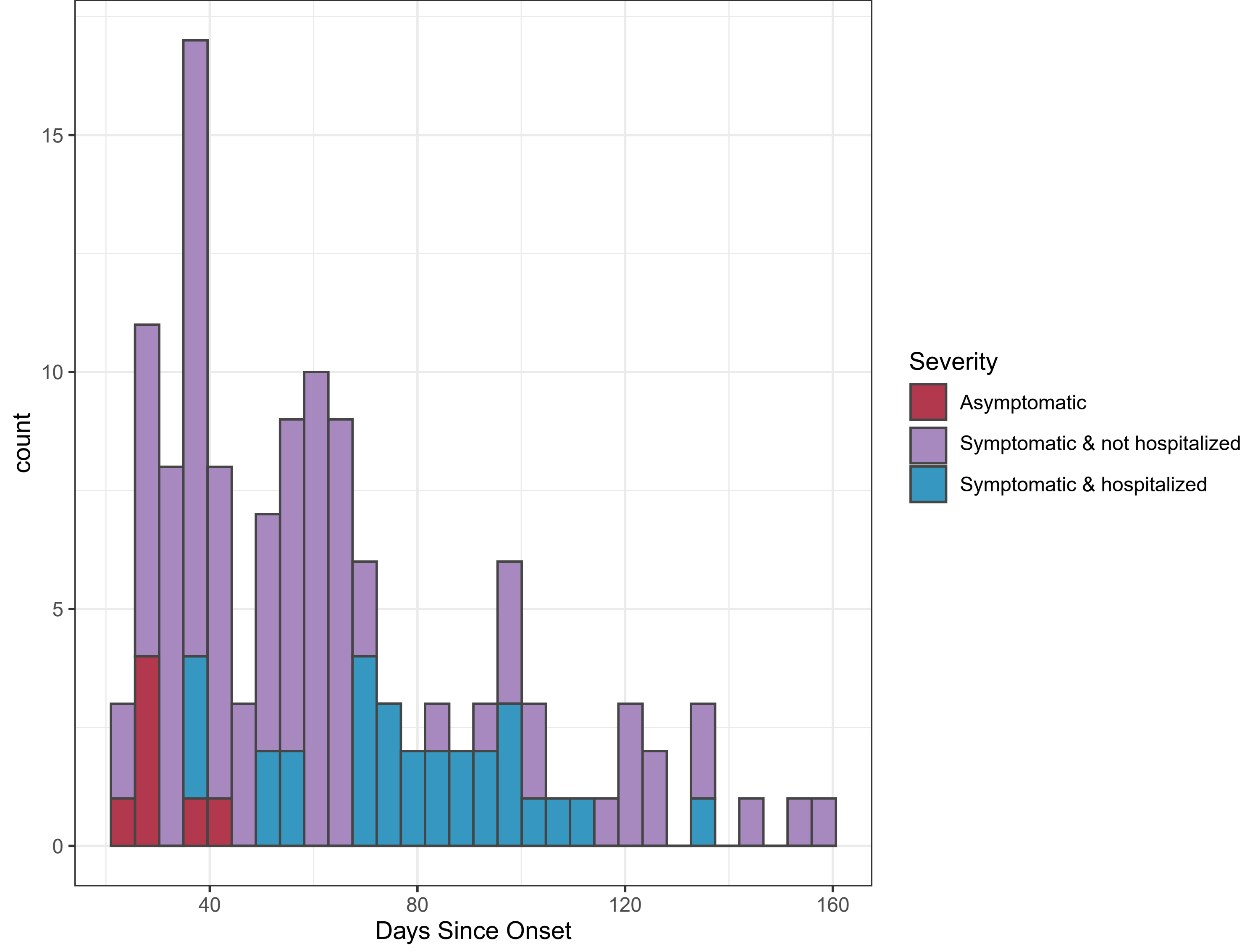

### Supplemental Figure 2

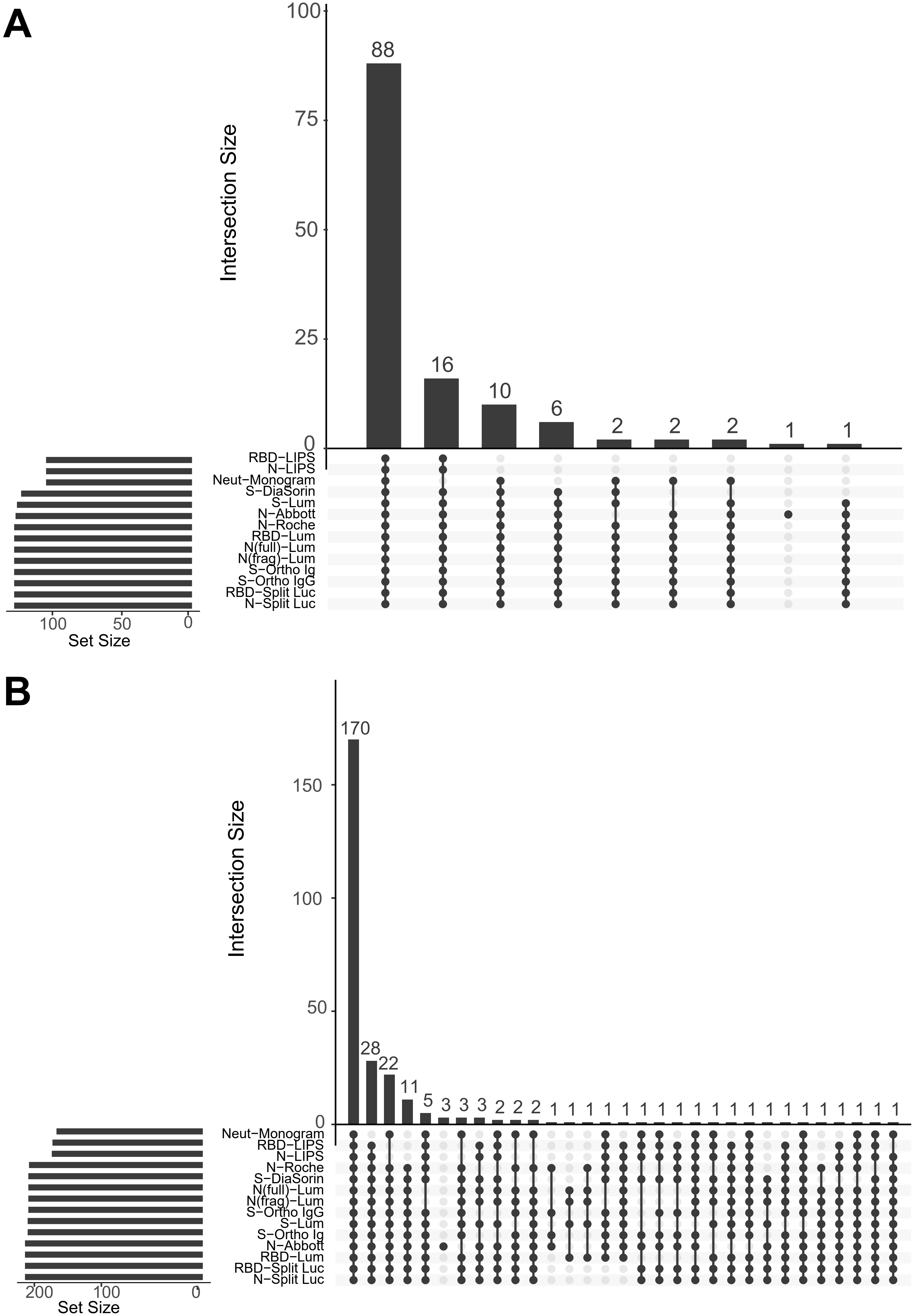

### Supplemental Figure 3A

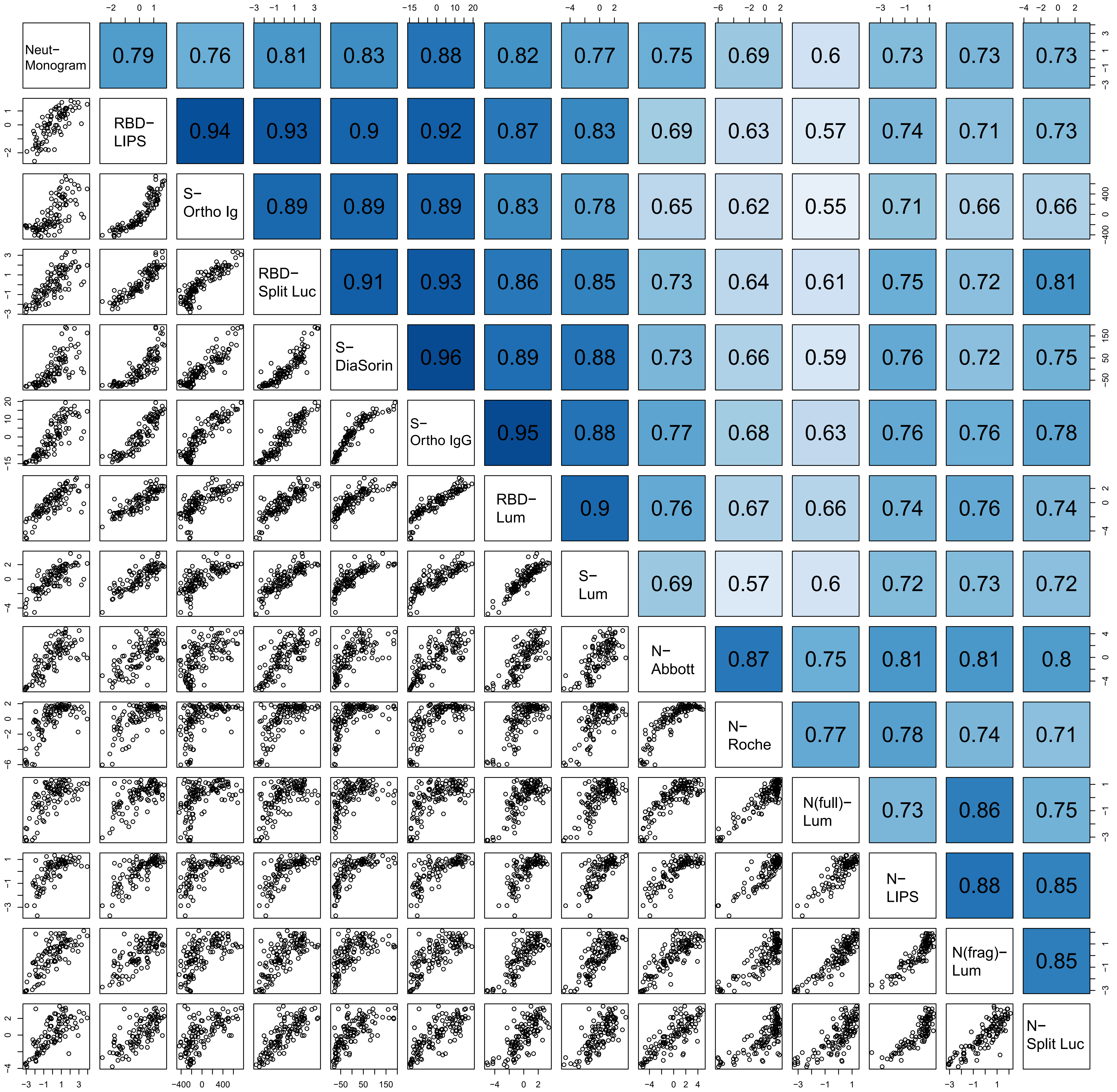

### Supplemental Figure 3B

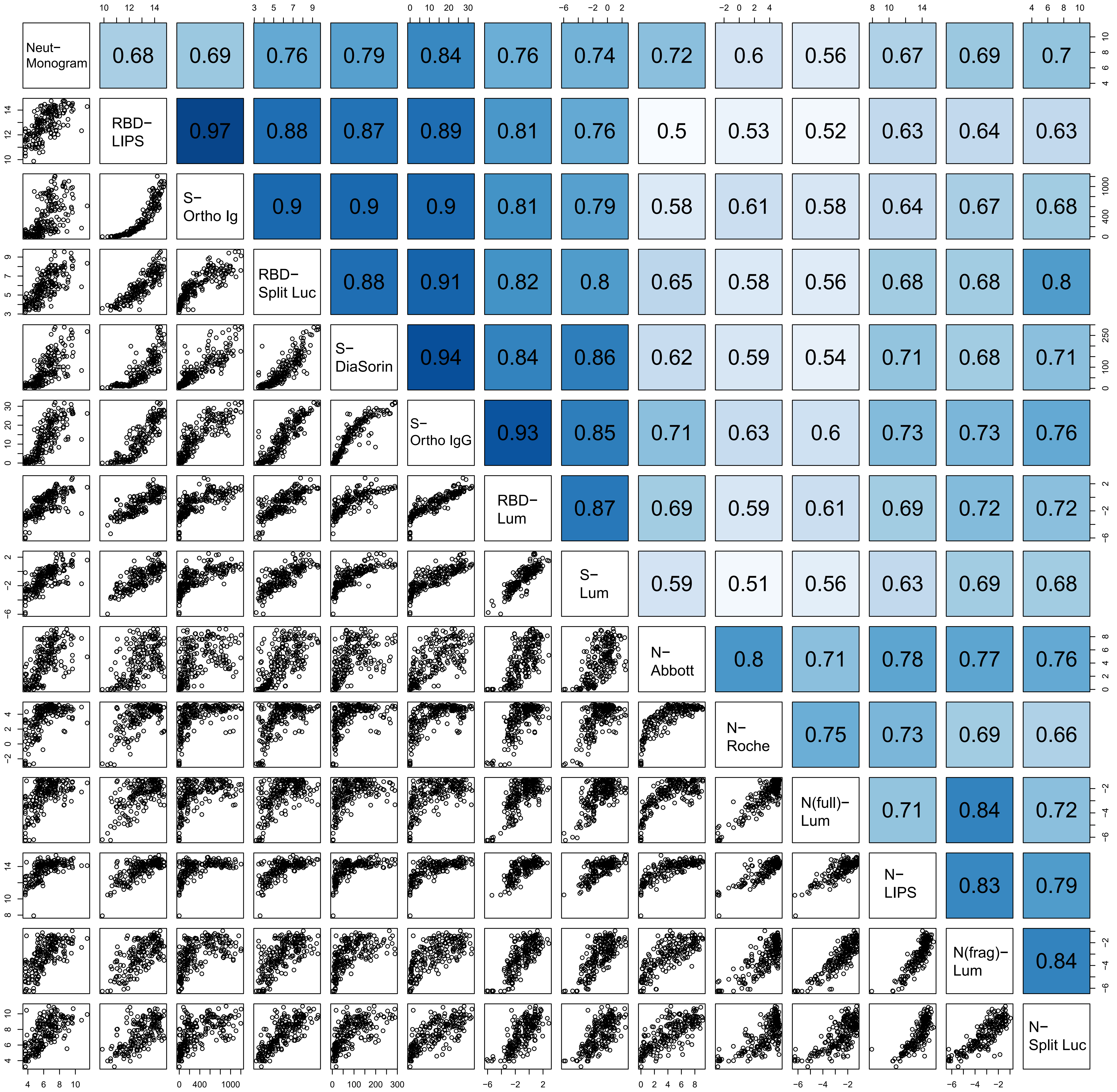

### Supplemental Figure 3C

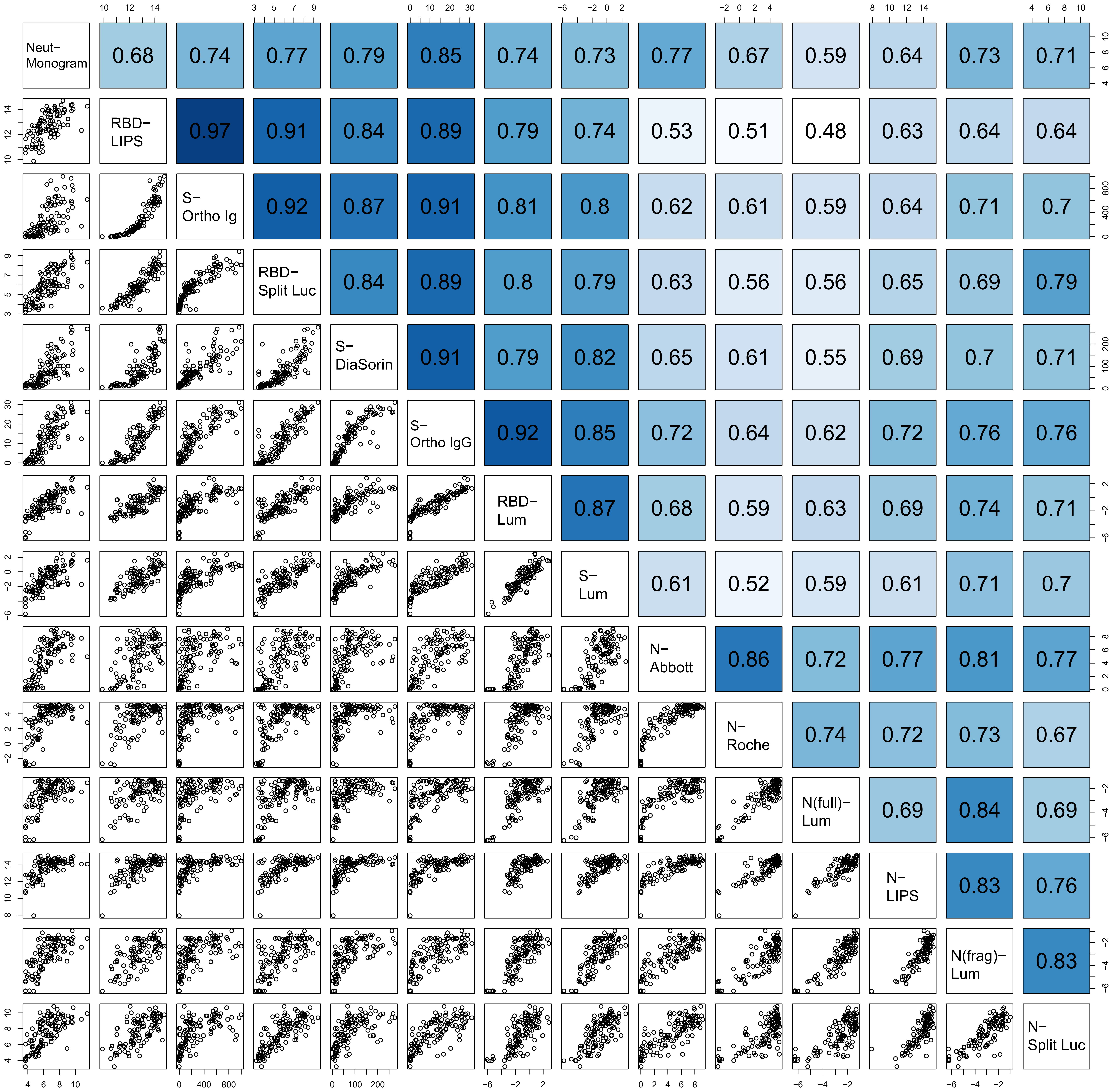

### Supplemental Figure 3D

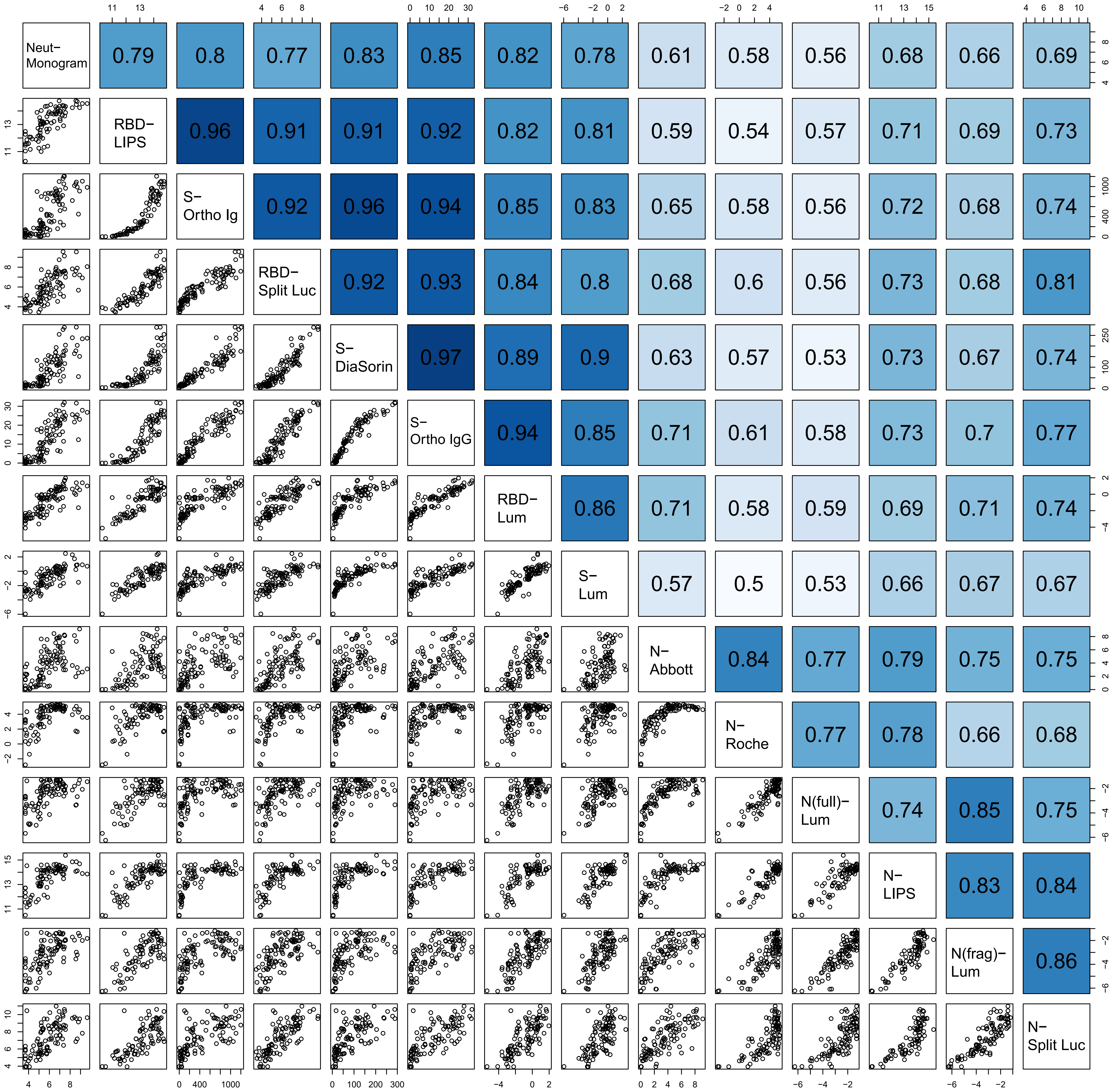

### Supplemental Figure 4

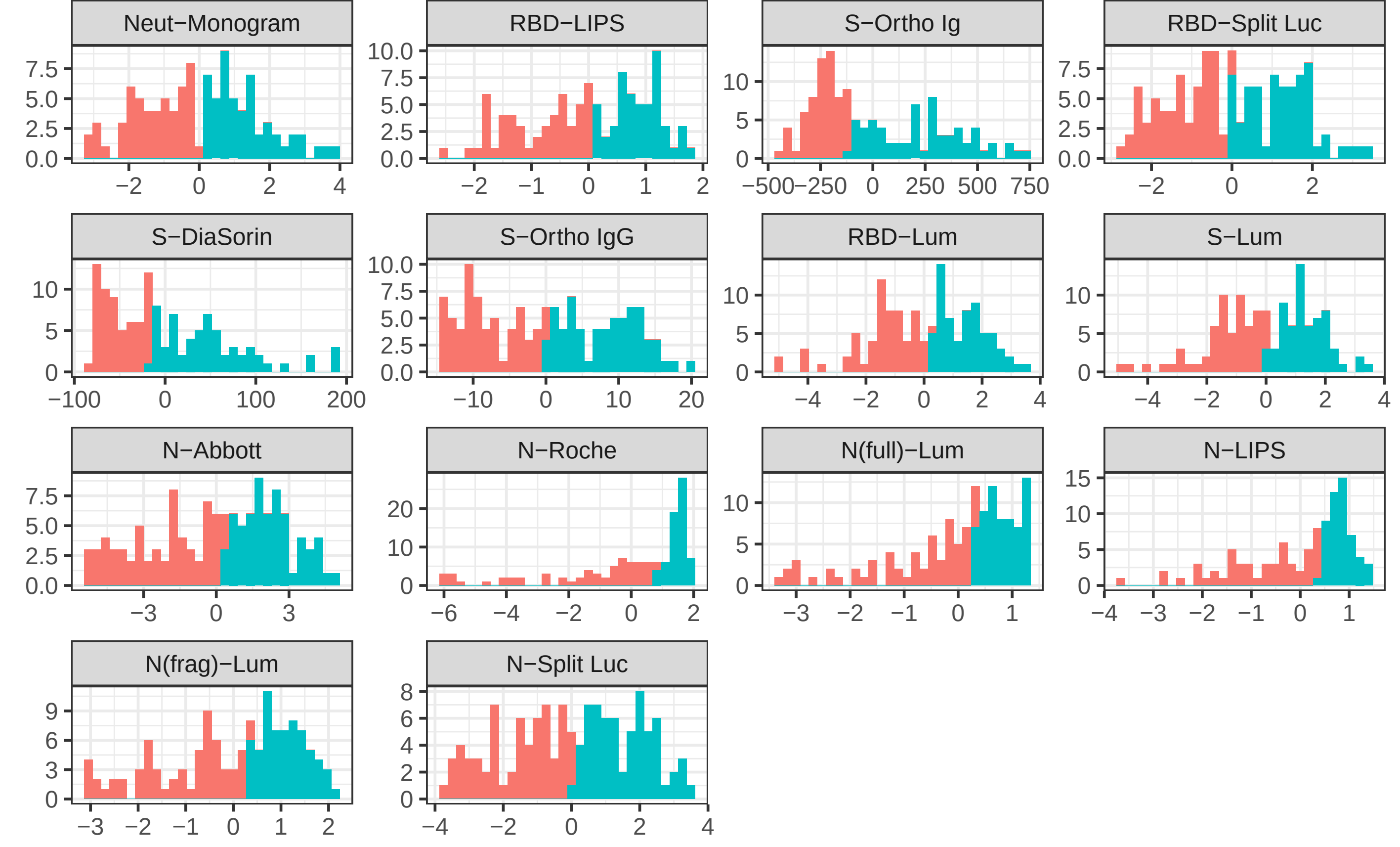

### Supplemental Figure 5

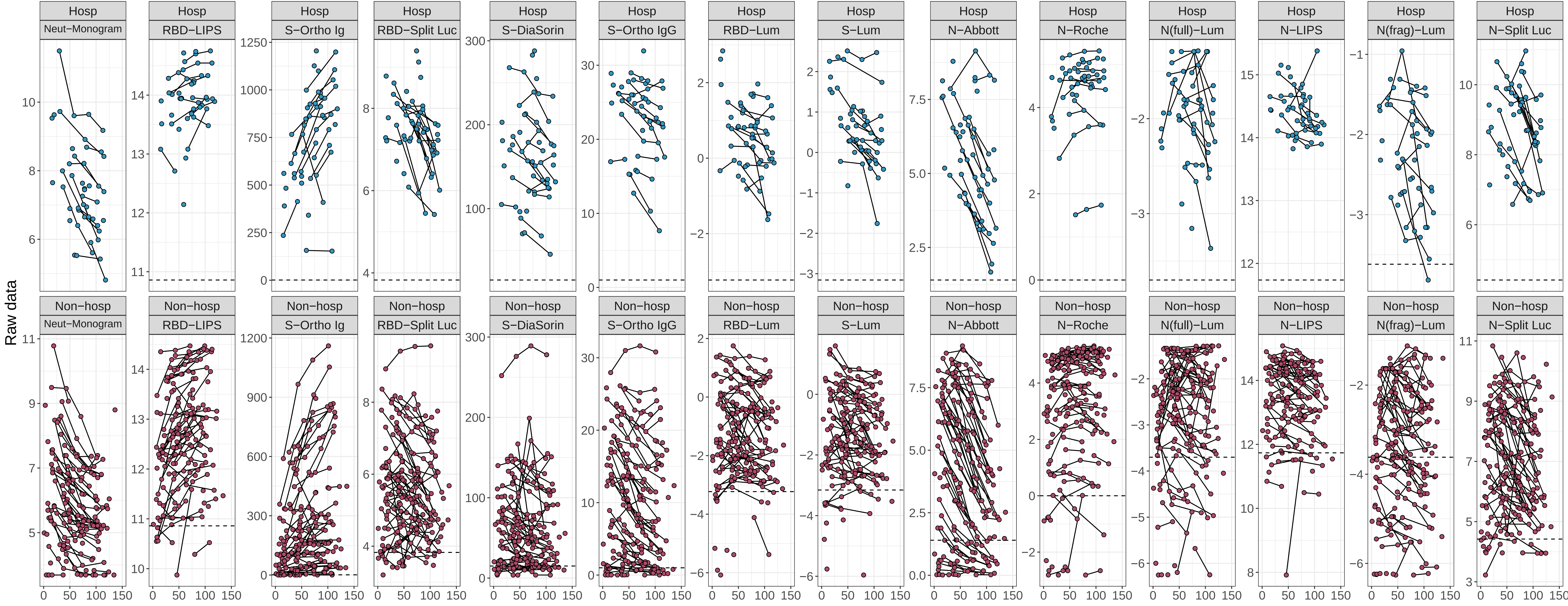

### Supplemental Figure 6

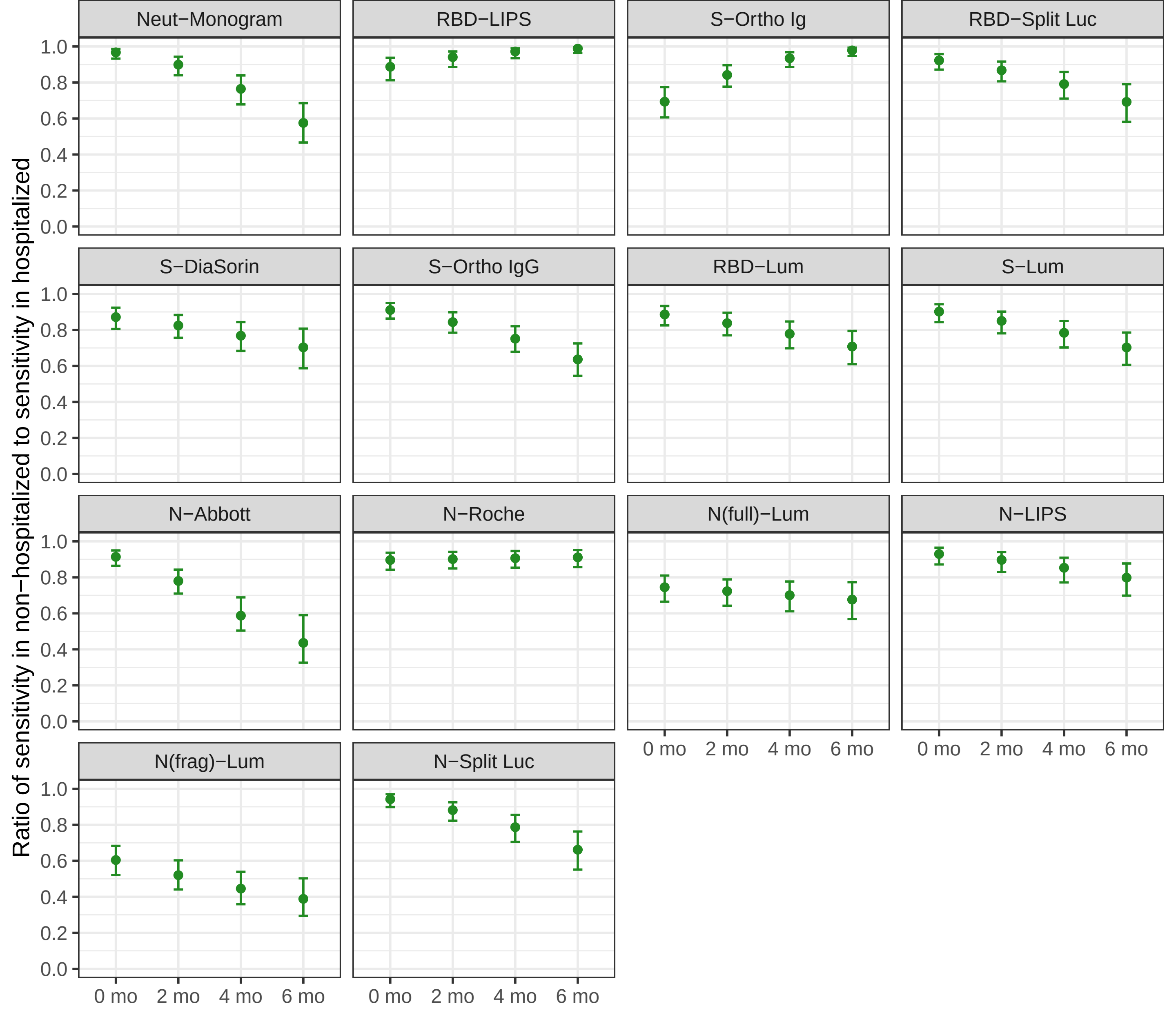

### Supplemental Figure 7

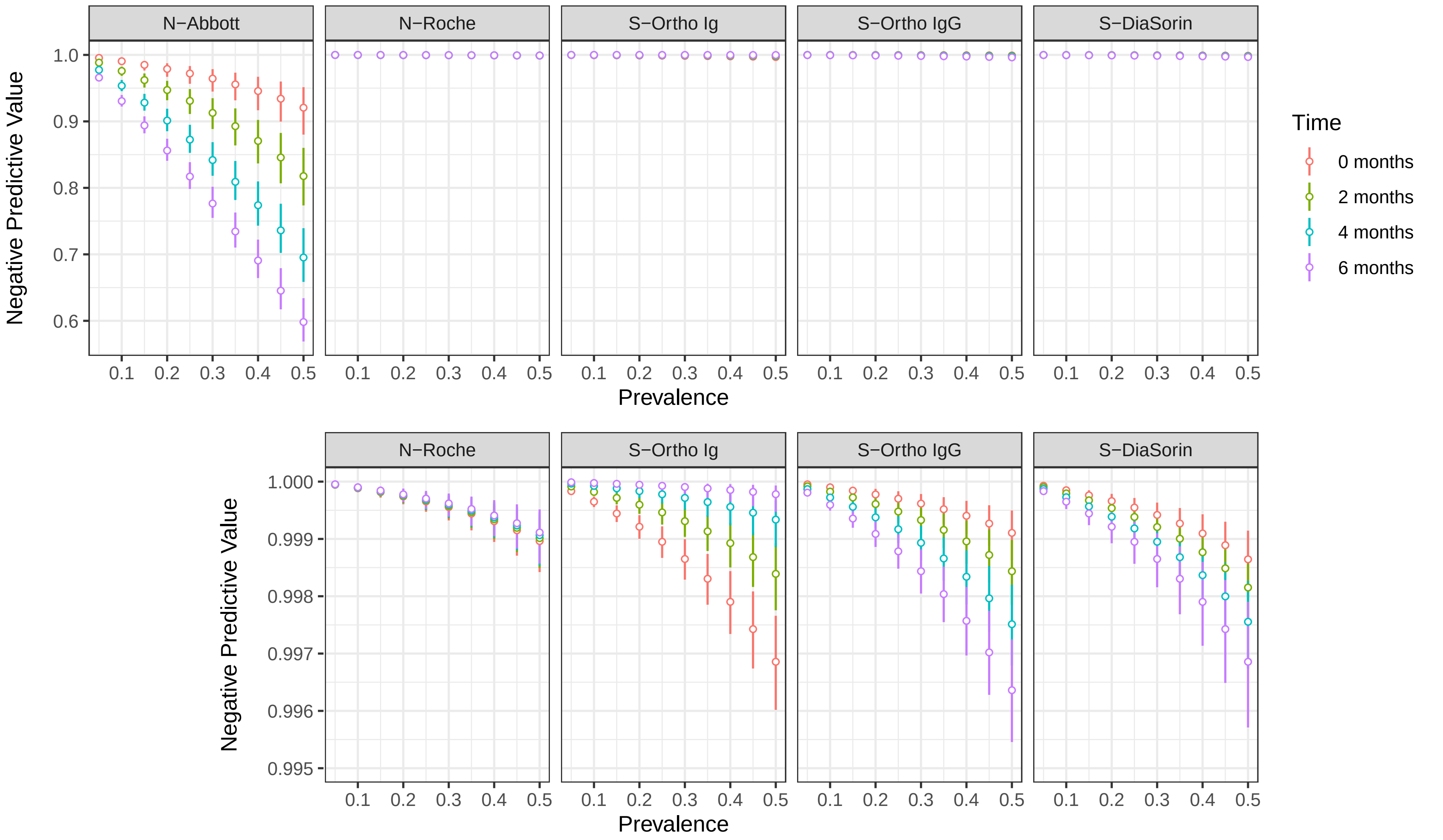
